# A Modified Delphi Consensus-based Comprehensive Checklist and Angoff Standard for Assessment of Competency in Brain Death/Death by Neurologic Criteria Determination

**DOI:** 10.1101/2025.08.27.25334589

**Authors:** Daniel S. Harrison, Neil Dhruva, Jenna L. Ford, David M. Greer, Sarah Wahlster, Nikita Chhabra, Nicholas A. Morris, the SECOND Study Group

## Abstract

**Objective:** To develop a comprehensive checklist, define critical actions, and establish a minimal passing standard for adult and pediatric critical care clinicians as well as other providers to facilitate formative and summative assessment of brain death/death by neurologic criteria (BD/DNC) determination Design: A pre-specified three round modified Delphi consensus process to define checklist items followed by a modified Angoff standard setting process to determine critical actions and item average ratings.

**Setting:** Electronic surveys.

**Subjects:** Selected authors of the 2023 Pediatric and Adult Brain Death/Death by Neurologic Criteria Consensus Practice Guideline, World Brain Death Project, and experts recommended by these authors (n=16) participated in the Delphi panel. Neurocritical care United Council for Neurologic Subspecialties and Accreditation Council for Graduate Medical Education examination committee members (n= 13) participated in Angoff standard setting.

**Interventions:** None.

**Measurements and Main Results:** A total of 98 unique checklist items related to assessment of prerequisites (23 items), performance of the clinical examination (28 items), apnea testing (36 items), and ancillary testing (11 items) were retained by the Delphi panel. Seven items were designated as critical actions based upon Angoff panelist consensus. The remaining 91 items were assigned item average ratings. The minimum passing score for an assessment including all items was set at 89%.

**Conclusions:** These guideline-concordant consensus checklist items, including critical actions and non-critical actions with their assigned item average ratings, may be applied selectively to simulated cases of BD/DNC determination for adults and children to determine a minimum passing score and readiness for independent practice, mitigating risk of inaccurate BD/DNC determination among critical care clinicians. Our process for systematically defining critical actions on a behavior checklist may be replicated for simulation-based summative assessment of learners in other critical care scenarios.

**KEY POINTS:** *Question:* What behaviors should be included in a checklist for assessment of brain death/death by neurologic criteria (BD/DNC) determination and what level of performance is consistent with readiness for independent practice?

*Findings:* A modified Delphi panel identified 98 checklist items which may be applied selectively to simulated cases of BD/DNC. An Angoff panel identified 7 critical actions and set average ratings for all other items.

*Meaning:* These guideline-concordant consensus checklist items and their average ratings may be applied to assessments of simulated or real cases of BD/DNC, including those intended for summative assessment, to determine learner readiness for independent determination.

## INTRODUCTION

The 2023 Pediatric and Adult Brain Death/Death by Neurologic Criteria (BD/DNC) Consensus Guideline highlights intensivists among “adequately trained and competent [clinicians] in the evaluation of BD/DNC in children or adults (1).” However, exposure to BD/DNC determination during training is inconsistent and there is significant variability in individual competence in BD/DNC determination, suggesting an ongoing need for optimization of education and assessment practices (2, 3). BD/DNC determination poses a unique challenge for educators in critical care as determination should be 100% accurate for this vastly important diagnosis. Those certifying learners for independent practice must have the highest level of certainty that these individuals are competent to determine BD/DNC. While written examinations are effective in assessing learner knowledge, knowledge does not directly translate into real-life performance (4–6). Simulation-based mastery learning (SBML) is a powerful educational tool that produces “excellence without exception”, a goal that aligns well with BD/DNC determination (7). SBML leads to improved learner performance in multiple critical care domains, including ventilator management (8), emergency response (9), communication (10), and critical care procedural skills (11, 12), improving patient outcomes in this context. SBML is a competency-based approach that relies on assessment and achievement of a minimal passing score.

Previously published simulation-based assessments of BD/DNC determination competence generally lack methodological rigor, including assessment and report of passing standards (2, 13, 14). The Pediatric and Adult BD/DNC Consensus Guideline includes a checklist to assist with clinical determination, but it was not designed for assessment and does not specify how items on the checklist should be completed (1). A standardized assessment with validity evidence for identifying a clinician demonstrating the minimum competence required for independent BD/DNC determination as part of a SBML curriculum has not been described. As such, we aimed to develop a comprehensive checklist, define critical actions, and set a minimal passing standard to facilitate formative and summative assessment of BD/DNC determination.

## MATERIALS AND METHODS

A modified Delphi consensus process was applied to define checklist items. Thereafter, a modified Angoff process was employed to determine critical items and to determine a minimum passing score (Figure 1, 15). This procedure is generally accepted to establish defensible passing scores on examinations in healthcare education (16). All study instruments were completed online anonymously using Qualtrics. Panelists completed an instrument to collect information on their expertise in BD/DNC determination and medical education.

**Figure 1.**
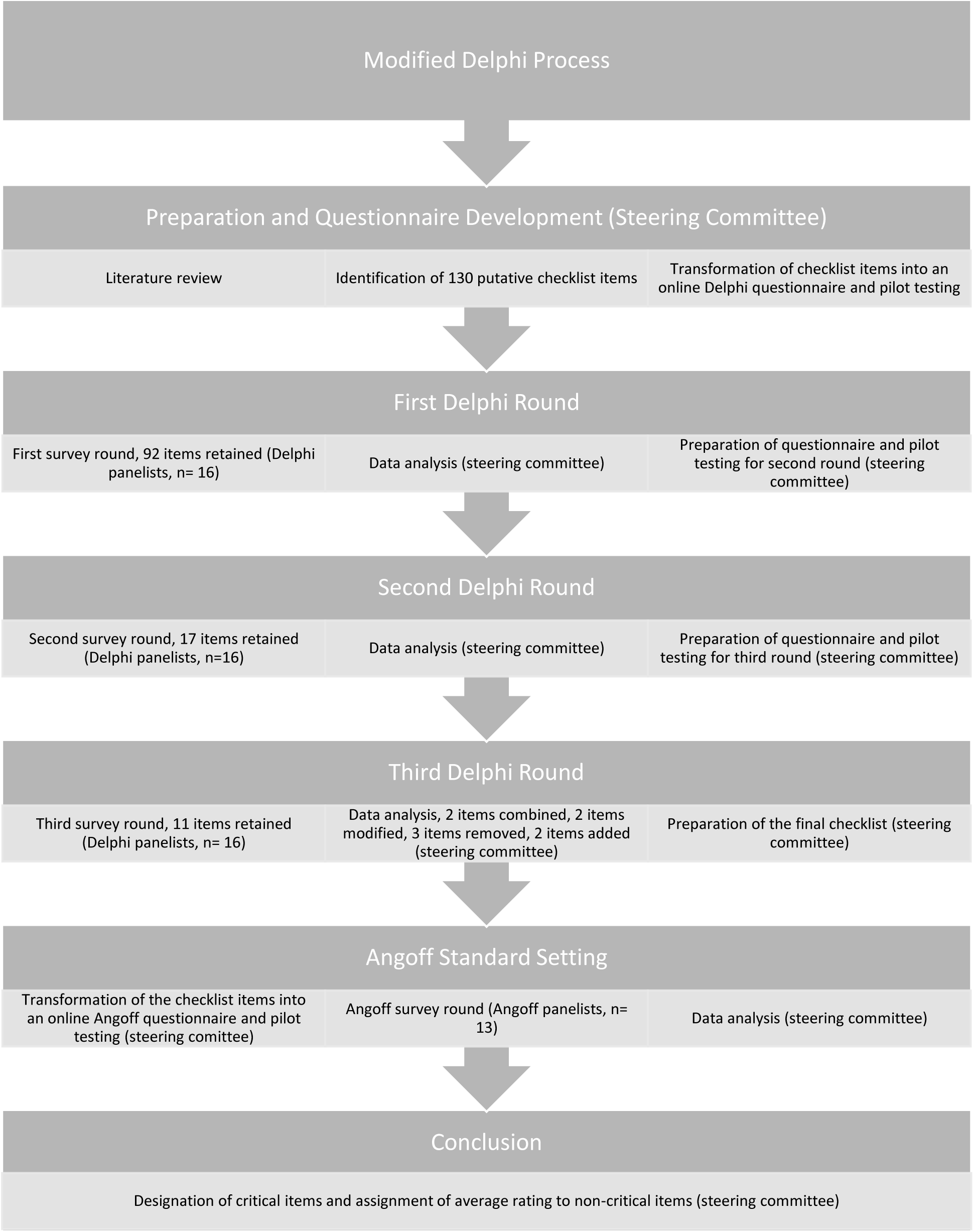
Study protocol

### Modified Delphi Consensus Process

Delphi panelists included selected authors of the Pediatric and Adult BD/DNC Consensus Guideline and World Brain Death Project, as well as those with national or international recognition as experts in BD/DNC determination referred by these authors (1, 17). The steering committee performed a literature review and a review of available checklists for BD/DNC determination, including from the Pediatric and Adult BD/DNC Consensus Guideline, and identified putative checklist items. These items were designed to be applied for learners either with or without required expertise in managing cardiopulmonary conditions (with the stipulation that learners without expertise in cardiopulmonary management must request assistance from a clinician with expertise for certain items during the apnea test). Items were transformed into a Delphi questionnaire. Before each Delphi round, the questionnaire was piloted and adjusted as needed by the steering committee.

Before beginning the Delphi process, a webinar was hosted to explain the rules and give Delphi panelists the opportunity to ask questions about the process. The rules were also included in writing at the beginning of the Delphi questionnaire itself. The steering committee pre-specified three Delphi rounds.

During round one, Delphi panelists were asked to vote how important it is that a learner complete specific actions during a simulated assessment of competency in independent BD/DNC determination on a 5-point scale (ranging from “not at all important” to “mandatory”) as previously described (18). Delphi panelists were instructed that all items that received greater than or equal to 80% "mandatory” or “very important" votes would be included in the behavior checklist. All items that received less than or equal to 20% "mandatory” or “very important" votes were excluded. All items that receive greater than 20% but less than 80% “mandatory” or “very important” votes underwent an additional round of voting. There is significant variability in the definition of consensus for Delphi studies, with thresholds typically ranging from 51-80% and some studies requiring as high as 97% agreement for item retention (19–21). A relatively rigorous threshold within the typical range (80%) was selected to limit the length of the checklist, anticipating a high level of agreement among the Delphi panelists.

Delphi panelists were allowed to make comments on and suggestions for each item during each round. During the first round, Delphi panelists were additionally allowed to suggest new items. After voting was complete, the steering committee met to review the data, discuss panelist comments, add new items, and modify existing items as suggested by the panel. Panelists were contacted individually as needed to clarify comments.

The panelists then completed a second round of voting. During the second and third rounds of voting, panelists had access to their individual votes from the prior round, as well as the average and standard deviation of the panel’s vote for each item. Panelists could not see individual votes of the other panelists. They were instructed that all new or modified items were subject to the same rules for retention, revoting, and rejection as in the first round. All unmodified items subject to revoting that received a "mandatory” or “very important" vote from greater than or equal to 80% of panelists were included in the behavior checklist. All unmodified items subject to revoting that received less than 80% "mandatory” or “very important" votes were excluded.

After the second round of voting was complete, the steering committee met to review the data, discuss panelist comments, and modify existing items as suggested by the panel. The panelists then completed a final round of voting. They were instructed that all items that received a "mandatory” or “very important" vote from greater than or equal to 80% of panelists would be included in the behavior checklist. All other items were excluded. The steering committee met a final time to review the data, discuss panelist comments, and certify the final list of checklist items.

### Modified Angoff Standard Setting with Critical Actions

Angoff panelists were members of the Accreditation Council for Graduate Medical Education and United Council for Neurologic Subspecialties neurocritical care (NCC) certification exam committees with publically available contact information. These individuals were selected because of their expertise in both critical care, including BD/DNC determination, and learner assessment.

Checklist items from the Delphi process were transformed into an Angoff questionnaire. The Angoff questionnaire was piloted and adjustments were made as needed by the steering committee. Before beginning the Angoff process, a webinar was hosted to explain the rules and give panelists the opportunity to ask questions. The rules were also included in writing at the beginning of the Angoff questionnaire.

Panelists were asked to indicate the percentage of minimally competent participants (individuals they would trust to make an independent BD/DNC determination) that they would expect to complete the specific actions during simulated assessment of BD/DNC. Given the very high stakes of BD/DNC determination, a separate mastery standard was not set because in this context the steering committee felt that the “minimally competent” candidate must be “well-prepared” to determine BD/DNC independently.

Similarly, the steering committee recognized that the Angoff method does not allow for systematic designation of behavior checklist items as truly critical (i.e. required for passing the assessment), but given the uniquely high stakes of BD/DNC determination the steering committee resolved that some items should receive such a designation. Other standard setting methods similarly do not allow for designation of critical actions (22, 23). The “Patient Safety” approach does discriminate between “essential” and “nonessential” checklist items and applies a conjunctive rule in which performance of nonessential items does not compensate for nonperformance of essential items (24). However, the Patient Safety approach allows participants who have missed one or more essential items to pass an assessment and is not consistently more stringent than the Angoff method (25). As such, we modified the Angoff method to allow for delineation of critical actions. Panelists were instructed that there would be some items they felt were critical (i.e. the participant automatically fails the assessment if they do not correctly complete the action). For these items, panelists were instructed to vote “100%”. Any item that received a 100% vote from more than 80% of the panel was designated as critical. Panelists were instructed that participants would be expected to complete all critical items and achieve the minimum passing score for the remaining checklist items to pass the assessment.

After voting, the steering committee met to review the data and identify outlier responses from panelists. Panelists were contacted as needed to confirm outlier responses. Once outlier responses were confirmed, the steering committee certified the final list of critical actions, item average ratings, and the overall minimal passing score.

### Statistics

All results are reported using descriptive statistics. All data was analyzed in Microsoft excel.

### Standard Protocol Approvals, Registrations, and Patient Consents

The study was exempt by the Boston University / Boston Medical Center institutional review board (“Simulation-Based Mastery Learning to Assess and Ensure Competency in Brain Death/Death by Neurologic Criteria Determination”, IRB number H-45585, approved 2/11/2025). The procedures followed were in accordance with the ethical standards of the responsible institutional committees on human experimentation and with the Helsinki Declaration of 1975, as most recently amended.

## RESULTS

Twenty experts were invited to participate in the Delphi panel and 16 accepted, including adult and pediatric clinicians. Delphi panelist qualifications are summarized in Table 1. There was no attrition of panelists between rounds.

**Table 1.** Delphi and Angoff panelist qualifications.

| Qualification | Delphi panel (n= 16) | Angoff panel (n= 13) |
| --- | --- | --- |
| Years in practice | 23.6 ± 11.6 | 18.5 ± 9.1 |
| Prior BD/DNC determinations | 201.6 ± 159.1 | 270 ± 338.4 |
| BD/DNC publications | 13.1 ± 18.7 | 1.2 ± 2.7 |
| Medical education publications | --- | 6.9 ± 9.2 |
| Years of BD/DNC teaching experience | 22.0 ± 10.3 | 16.1 ± 7.9 |
| Experience determining competency standards for clinical assessment | 12 (75.0%) | 11 (84.6%) |
| BD/DNC simulation teaching experience | 14 (87.5%) | 8 (61.5%) |
| Developed local BD/DNC protocol | 15 (93.8%) | 12 (92.3%) |
| Invited speaker on BD/DNC | 16 (100%) | 7 (53.8%) |
| National/international committee service in BD/DNC | 11 (68.8%) | 2 (15.4%) |
| Board Certification |  |  |
| Neurocritical Care | 11 (68.8%) | 13 (100%) |
| Adult Neurology | 10 (62.5%) | 5 (38.5%) |
| Pediatric Critical Care Medicine | 2 (12.5%) | 0 (0%) |
| Pediatric Neurology | 1 (6.3%) | 0 (0%) |
| Neurosurgery | 1 (6.3%) | 0 (0%) |
| Pediatrics | 1 (6.3%) | 0 (0%) |
| Critical Care Medicine | 1 (6.3%) | 1 (7.7%) |
| Internal Medicine | 1 (6.3%) | 1 (7.7%) |
| Critical Care EEG | 1 (6.3%) | 0 (0%) |
| Emergency Medicine | 0 (0%) | 2 (15.4%) |
| Anesthesiology | 0 (0%) | 2 (15.4%) |

BD/DNC= brain death/death by neurologic criteria

The steering committee developed an initial list of 130 putative checklist items, and Delphi panelists suggested an additional 8 checklist items (eAppendix 1). After round one of voting 92 items were retained, after round two 17 additional items were retained, and after round three 11 additional items were retained. Delphi panelists made over 500 suggestions for item modifications in rounds one and two, including suggestions for modifications to items which met criteria for retention. This resulted in modifications and revoting on some items which initially met consensus for retention. To avoid overlap and ensure consistency and clarity in the final checklist, the steering committee combined two items, modified two items, removed three items, and added two items (eAppendix 2). The steering committee did not add any items which were rejected by the panel. The final checklist included 98 unique items, related to assessment of prerequisites (23 items), performance of the clinical examination (28 items), apnea testing (36 items), and ancillary testing (11 items, eTable 1).

Twenty-two experts were invited to participate in the Angoff panel. Thirteen accepted and ultimately participated in the study. Angoff Panelist qualifications are summarized in Table 1. Seven items met the pre-specified 80% consensus threshold to be designated as “critical”. These items included identifying the mechanism of BD/DNC, assessing cranial nerve reflexes, checking an ABG before beginning the apnea test, and aborting the apnea test in a patient who demonstrates spontaneous respiratory effort (Table 2). Item average ratings for the remaining 91 items are available in eTable 1. The minimum passing score for an assessment including all items was set at 89%.

**Table 2.** Critical action items.

| Item | Percent agreement |
| --- | --- |
| Participant does NOT perform a clinical exam to determine BD/DNC if unable to confirm a catastrophic, permanent brain injury caused by an identified mechanism that is known to lead to BD/DNC. | 84.6% |
| Participant tests pupillary responses to bright light bilaterally. | 84.6% |
| Participant tests the corneal reflex bilaterally. | 84.6% |
| Participant assesses for a gag reflex. | 84.6% |
| Participant assesses for a cough reflex. | 84.6% |
| Participant requests baseline arterial blood gas. | 84.6% |
| If there is spontaneous respiratory effort, the participant reconnects the ventilator and does NOT continue with further BD/DNC evaluation. | 83.3% |

BD/DNC= brain death/death by neurologic criteria

## DISCUSSION

Using expert consensus and best practices for mastery learning, we created a guideline-concordant, comprehensive checklist for formative and summative assessment of BD/DNC determination. We additionally set a minimum passing score, which will facilitate determination of learner readiness for independent practice. Standardization of BD/DNC education and assessment may mitigate longstanding concerns about practice variations and minimize the risk of inaccurate BD/DNC determination among critical care clinicians.

While the complete checklist has more items than could be feasibly applied to a single simulated scenario, many checklist items are only applicable to some cases of BD/DNC, since they involve addressing various pre-requisites to testing (eTable 2). When applying the checklist to cases of simulated BD/DNC developed by the authors, resulting case-specific checklists, including cases with multiple confounders to BD/DNC determination, typically had 40-45 items each. This longer than average checklist is typical for critical care procedures (including determination of BD/DNC), in which many actions must be completed correctly to achieve an accurate result (8, 26). As not all checklist items will be tested in an individual case, when applying this checklist for assessment of competency in BD/DNC determination, we suggest incorporating several cases with different confounders for a more comprehensive assessment.

The steering committee identified significant variability and even inaccuracies in prior checklists for clinical BD/DNC determination, consistent with observed variability in physician and institutional practices in BD/DNC determination (27–29). For example, one unpublished checklist available to the authors included an item for checking a jaw jerk reflex and another suggested that pupil size smaller than 4mm was inconsistent with BD/DNC. While the Pediatric and Adult BD/DNC Consensus Guideline provides a checklist for clinical determination, it was not developed to assess competency. Thus, it lacks specificity regarding how to perform key components of the clinical examination and apnea test. In contrast, the guideline concordant checklist developed herein was developed specifically for the purpose of competency assessment, assessing both item completion and proper technique, as defined by an expert panel. We expect our checklist to help standardize BD/DNC determination competency assessment, with the ultimate goal of reducing variability in BD/DNC determination (30).

In SBML, checklist development is typically followed by Angoff standard setting to determine a mastery score (15). Though a Delphi panel has determined that a group of items belong on a checklist, this does not imply that all items are of equal importance. In fact, some actions may be so critical that a participant should not pass the assessment without completing the item. Our methodology, which borrows from the Angoff and Patient Safety standard setting approaches, could be applied for assessment of learners in other critical care scenarios (for example, cardiac arrest resuscitation) where some items are so critical to patient outcomes (for example, defibrillating a patient with a shockable rhythm) that their omission should automatically lead to a failure of the assessment.

Using this method, there were items which surprisingly did not meet the threshold for inclusion as critical actions including checking the oculovestibular reflex (OVR) and declaring BD/DNC if the exam, apnea test, and ancillary test (when indicated) are consistent with BD/DNC. While there were possible explanations for why panelists may have considered these items non-critical, this led us to consider whether the threshold for considering an item to be critical was too high and should be lowered or whether the procedure should be altered (e.g. subjecting items to a second round of voting with feedback on the votes of the rest of the panel, as in the Delphi process). Notably, these items are critical actions for assessing competency in BD/DNC determination (as opposed to critical actions in the determination of BD/DNC itself). In applying the checklist to three unique cases of simulated BD/DNC determination developed by the authors, the resulting minimum passing standard (100% completion of 5 critical actions and 89% completion of all non-critical items) was consistent with or more rigorous than the mastery standard in other examples of simulation-based assessment (8, 26, 31). As such, the *a priori* definition was maintained.

The checklist includes items of omission (those which a participant should NOT perform), which are generally avoided in assessment. Several alternatives were considered and experts were consulted for opinions regarding how to best handle these items. For example, whether the participant should be required to verbalize their actions throughout the exam was considered. However, this would limit the response process validity of the assessment. Whether participants should be required to write a BD/DNC determination note after completing the simulated assessment was also considered. However, not all simulated assessments for BD/DNC will include a note, which would limit the generalizability of the checklist. Ultimately, items of omission were maintained given the potential pitfalls of the alternative strategies.

This study has several limitations. Delphi and Angoff panelists were from the United States and Canada, but BD/DNC policies differ among institutions and countries (32, 33). Though the checklist developed here is concordant with the Adult and Pediatric BD/DNC Consensus Guideline, it is not expected to be concordant with the local policy at every institution or in every country and may not account for regional or cultural practices. The checklist is longer than average for summative assessment in SBML—this could be burdensome for raters. The rationale for the chosen consensus threshold is described above, however, a different threshold may have resulted in a different number of critical actions and minimal passing score in some scenarios. Traditional Angoff standard setting language (as opposed to mastery Angoff language) was used to determine the item average ratings. Though the minimally competent candidate was defined as one ready for independent practice, thresholds developed using these item average ratings will be minimum passing scores rather than mastery standards. No case to which the checklist is applied could realistically include all possible checklist items. As such, the cases developed for a given assessment will shape their case-specific checklists, which could introduce variability between assessments. In practice, this can be addressed by using multiple cases in an assessment or by adding a complementary knowledge assessment that focuses more on cognitive skills of ruling out confounders, such as that offered as part of the Neurocritical Care Society’s online Brain Death Determination course (34).

In the future, this checklist can be applied to SBML curricula for BD/DNC determination. It could also be applied for learner assessment in real clinical settings on actual patients. The checklist could further be applied for individual certification of BD/DNC determination competence. While this checklist includes technical skills required for independent BD/DNC determination, a parallel checklist to assess for BD/DNC determination-related communication skills should be developed.

## CONCLUSIONS

These guideline-concordant consensus checklist items and minimal passing standard may be applied to simulated or real cases of BD/DNC to assess competency in BD/DNC determination.

## Supporting information

eAppendix 1

eAppendix 2

eTable 1

eTable 2

## Data Availability

All data produced in the present study are available upon reasonable request to the authors.

## ACKNOWLEDGEMENTS

The authors thank Drs. Edward Krupat, Demian Syzyld, and William McGaghie for their input in checklist development and standard setting methodology.

## REFERENCES

1. Greer DM, Kirschen MP, Lewis A, et al.: Pediatric and adult brain death/death by neurologic criteria consensus guideline: report of the AAN Guidelines Subcommittee, AAP, CNS, and SCCM. Neurology 2023; 101:1112–1132

2. MacDougall BJ, Robinson JD, Kappus L, et al.: Simulation-Based Training in Brain Death Determination [Internet]. Neurocrit Care 2014; 21:383–391Available from: 10.1007/s12028-014-9975-x

3. Ausmus AM, Simpson PM, Zhang L, et al.: A Needs Assessment of Brain Death Education in Pediatric Critical Care Medicine Fellowships* [Internet]. Pediatric Critical Care Medicine 2018; 19Available from: https://journals.lww.com/pccmjournal/fulltext/2018/07000/a_needs_assessment_of_brain_dea th_education_in.6.aspx

4. Wayne DB, John B, Viva J. S, et al.: Simulation-Based Training of Internal Medicine Residents in Advanced Cardiac Life Support Protocols: A Randomized Trial [Internet]. Teach Learn Med 2005; 17:202–208Available from: 10.1207/s15328015tlm1703_3

5. Wayne DB, Butter J, Siddall VJ, et al.: Mastery learning of advanced cardiac life support skills by internal medicine residents using simulation technology and deliberate practice. J Gen Intern Med 2006; 21:251–256

6. Wayne DB, Barsuk JH, O’Leary KJ, et al.: Mastery learning of thoracentesis skills by internal medicine residents using simulation technology and deliberate practice [Internet]. J Hosp Med 2008; 3:48–54Available from: 10.1002/jhm.268

7. McGaghie WC, Harris IB: Learning Theory Foundations of Simulation-Based Mastery Learning [Internet]. Simulation in Healthcare 2018; 13Available from: https://journals.lww.com/simulationinhealthcare/fulltext/2018/06001/learning_theory_foundati ons_of_simulation_based.3.aspx

8. Schroedl CJ, Frogameni A, Barsuk JH, et al.: Impact of Simulation-based Mastery Learning on Resident Skill Managing Mechanical Ventilators [Internet]. ATS Sch 2020; 2:34–48Available from: 10.34197/ats-scholar.2020-0023OC

9. Barsuk JH, Cohen ER, Wayne DB, et al.: Developing a Simulation-Based Mastery Learning Curriculum: Lessons From 11 Years of Advanced Cardiac Life Support [Internet]. Simulation in Healthcare 2016; 11Available from: https://journals.lww.com/simulationinhealthcare/fulltext/2016/02000/developing_a_simulation_based_mastery_learning.7.aspx

10. Smith MM, Walter JM, Munger NK, et al.: Excellence for All: Simulation-Based Mastery Learning for ICU Goals of Care Conversations [Internet]. Chest 2025; Available from: 10.1016/j.chest.2025.06.004

11. Barsuk JH, Cohen ER, Potts S, et al.: Dissemination of a simulation-based mastery learning intervention reduces central line-associated bloodstream infections [Internet]. BMJ Quality & Safety 2014; 23:749Available from: http://qualitysafety.bmj.com/content/23/9/749.abstract

12. Barsuk JH, Cohen ER, Williams M V, et al.: Simulation-Based Mastery Learning for Thoracentesis Skills Improves Patient Outcomes: A Randomized Trial [Internet]. Academic Medicine 2018; 93Available from: https://journals.lww.com/academicmedicine/fulltext/2018/05000/simulation_based_mastery_le arning_for.35.aspx

13. Hocker S, Schumacher D, Mandrekar J, et al.: Testing Confounders in Brain Death Determination: A New Simulation Model [Internet]. Neurocrit Care 2015; 23:401–408Available from: 10.1007/s12028-015-0130-0

14. Wasserstrom B, Maciel C, Wilson C, et al.: Simulation-based Assessment of Brain Death Determination for ACGME Milestones of Adult Neurology Residents (P8-2.004) [Internet]. Neurology 2023; 100:4578Available from: 10.1212/WNL.0000000000204078

15. Klein M, Loke D, Barsuk J, et al.: Twelve tips for developing simulation-based mastery learning clinical skills checklists. Med Teach 2024; 47:1–6

16. Downing SM, Tekian A, Yudkowsky R: RESEARCH METHODOLOGY: Procedures for Establishing Defensible Absolute Passing Scores on Performance Examinations in Health Professions Education [Internet]. Teach Learn Med 2006; 18:50–57Available from: 10.1207/s15328015tlm1801_11

17. Greer DM, Shemie SD, Lewis A, et al.: Determination of Brain Death/Death by Neurologic Criteria: The World Brain Death Project [Internet]. JAMA 2020; 324:1078–1097Available from: 10.1001/jama.2020.11586

18. Harrison DS, Sigman EJ, Ch’ang JH, et al.: A Modified Delphi Consensus Approach to Define Entrustable Professional Activities for Neurocritical Care Advanced Practice Providers. Crit Care Med 2024;

19. Diamond IR, Grant RC, Feldman BM, et al.: Defining consensus: A systematic review recommends methodologic criteria for reporting of Delphi studies [Internet]. J Clin Epidemiol 2014; 67:401– 409Available from: https://www.sciencedirect.com/science/article/pii/S0895435613005076

20. Hasson F, Keeney S, McKenna H: Research guidelines for the Delphi survey technique [Internet]. J Adv Nurs 2000; 32:1008–1015Available from: 10.1046/j.1365-2648.2000.t01-1-01567.x

21. Humphrey-Murto S, Lara V, Carol G, et al.: Using consensus group methods such as Delphi and Nominal Group in medical education research* [Internet]. Med Teach 2017; 39:14–19Available from: 10.1080/0142159X.2017.1245856

22. Ebel RL, Frisbie DA: Essentials of educational measurement. 1972;

23. Hofstee WKB: The case for compromise in educational selection and grading. In: Anderson S, editor(s). On educational testing. San Francisco: Jossey-Bass; 1983. p. 107–127.

24. Yudkowsky R, Tumuluru S, Casey P, et al.: A Patient Safety Approach to Setting Pass/Fail Standards for Basic Procedural Skills Checklists. Simul Healthc 2014; 9

25. Barsuk JH, Cohen ER, Wayne DB, et al.: A Comparison of Approaches for Mastery Learning Standard Setting [Internet]. Academic Medicine 2018; 93Available from: https://journals.lww.com/academicmedicine/fulltext/2018/07000/a_comparison_of_approaches_for_mastery_learning.33.aspx

26. Anderson K, Wang X, Batchelor T, et al.: 271 Transesophageal Echocardiography in the Emergency Department: Development of a Validated Checklist for Training [Internet]. Ann Emerg Med 2024; 84:S126Available from: 10.1016/j.annemergmed.2024.08.566

27. Braksick SA, Robinson CP, Gronseth GS, et al.: Variability in reported physician practices for brain death determination [Internet]. Neurology 2019; 92:e888–e894Available from: 10.1212/WNL.0000000000007009

28. Greer DM, Varelas PN, Haque S, et al.: Variability of brain death determination guidelines in leading US neurologic institutions [Internet]. Neurology 2008; 70:284–289Available from: 10.1212/01.wnl.0000296278.59487.c2

29. Francoeur C, Weiss MJ, MacDonald JM, et al.: Variability in Pediatric Brain Death Determination Protocols in the United States [Internet]. Neurology 2021; 97:e310–e319Available from: 10.1212/WNL.0000000000012225

30. Barnes E, Greer D: Inconsistency in Brain Death Determination Should Not Be Tolerated. AMA J Ethics 2020; 22:E1027–1032

31. Mikhaeil-Demo Y, Barsuk JH, Culler GW, et al.: Use of a simulation-based mastery learning curriculum for neurology residents to improve the identification and management of status epilepticus [Internet]. Epilepsy & Behavior 2020; 111:107247Available from: https://www.sciencedirect.com/science/article/pii/S1525505020304261

32. Lewis A, Bakkar A, Kreiger-Benson E, et al.: Determination of death by neurologic criteria around the world [Internet]. Neurology 2020; 95:e299–e309Available from: 10.1212/WNL.0000000000009888

33. Wahlster S, Wijdicks EFM, Patel P V, et al.: Brain death declaration [Internet]. Neurology 2015; 84:1870–1879Available from: 10.1212/WNL.0000000000001540

34. Rubin MA, Kirschen MP, Lewis A: The Neurocritical Care Brain Death Determination Course: Purpose, Design, and Early Findings [Internet]. Neurocrit Care 2021; 35:913–915Available from: 10.1007/s12028-021-01275-4

