## Supplementary material for "A Modified Delphi Consensus-based Comprehensive Checklist and Angoff Standard for Assessment of Competency in Brain Death/Death by Neurologic Criteria Determination": eAppendix 1

**eAppendix 1- Putative Checklist Items**

*Steering Committee Proposed*

1. If patient has NOT sustained a catastrophic, permanent brain injury caused by an identified mechanism that is known to lead to brain death/death by neurologic criteria, participant does NOT perform a clinical exam to determine BD/DNC .
2. Participant reviews neuroimaging studies.
3. If neuroimaging is inconsistent with mechanism and severity of brain injury or does NOT demonstrate catastrophic, permanent supratentorial injury, then participant does NOT perform a clinical exam to determine BD/DNC.
4. If neuroimaging is inconsistent with cerebral circulatory arrest, then participant does NOT perform the clinical exam to determine BD/DNC.
5. If insufficient time has passed since injury to ensure there is no potential for recovery of brain function as determined by the evaluator based on the pathophysiology of the brain injury (or <24h following an anoxic injury), then participant does NOT perform a clinical exam to determine BD/DNC.
6. If patient core body temperature is or has been ≤ 35.5, participant requests rewarming to ≥36C for ≥24 hours before performing a clinical exam to determine BD/DNC.
7. If SBP <100 (in a patient NOT on veno-arterial ECMO) or MAP <75, participant asks for BP augmentation before performing (or continuing with) a clinical exam to determine BD/DNC.
8. Participant asks about or reviews recently administered medications.
9. If a paralytic HAS been given, participant uses train-of-four stimulator or demonstrates deep tendon reflexes to exclude ongoing paralysis.
10. If DTRs are absent or there are no twitches on TOF, then participant asks to wait and reassess until one or both of these return before performing a clinical exam to determine BD/DNC.

11. Participant asks about or reviews drug levels, as available, for medications that may suppress central nervous system function.
12. If drug levels for medications that may suppress central nervous system function are supratherapeutic, pentobarbital level is >5mcg/mL, blood or urine toxicology screen is positive, or blood alcohol level is >80m/dL, then participant asks to recheck the level or screen after a delay before performing a clinical exam to determine BD/DNC.
13. If the patient received CNS-suppressing medications for which levels are NOT available, the participant asks to wait at least five half-lives for these drugs to have passed before performing a clinical exam to determine BD/DNC.
14. If the patient received CNS-suppressing medications for which levels are NOT available, the participant asks for a pharmacy consult (as available).
15. If the patient received CNS-suppressing medications for which levels are NOT available and there is renal/hepatic dysfunction or if the patient is obese or was hypothermic, the participant asks to wait longer than five half-lives before performing a clinical exam to determine BD/DNC.
16. If auto-triggering/cycling on the ventilator is suspected, then participant adjusts or requests adjustment to trigger sensitivity on ventilator.
17. The participant reviews or requests results of recent laboratory values.
18. If a severe metabolic, acid-base, or endocrine derangement is present and correctable, the participant proposes a specific work up and management plan to correct (or requests this be done by the primary team).
19. If the ammonia level is >75 µmol/L, the participant proposes a specific work up and management plan to correct (or requests this be done by the primary team).
20. If the blood urea nitrogen >75 mg/dL, the participant proposes a specific work up and management plan to correct (or requests this be done by the primary team).

© 2025 Daniel S. Harrison, Neil Dhruva, Jenna L. Ford, David M. Greer, Sarah Wahlster, Nikita Chhabra, Nicholas A. Morris

21. If the calcium (or ionized calcium) level is <7 or >11 mg/dL (or <1 or >1.3 mmol/L), the participant proposes a specific work up and management plan to correct (or requests this be done by the primary team).
22. If the glucose level is <70 or >300 mg/dL, the participant proposes a specific work up and management plan to correct (or requests this be done by the primary team).
23. If the magnesium level is <1.5 or >4 mg/dL, the participant proposes a specific work up and management plan to correct (or requests this be done by the primary team).
24. If the potassium level is <3 or >6 mmol/L, the participant proposes a specific work up and management plan to correct (or requests this be done by the primary team).
25. If the sodium level is <130 or >160 mmol/L, the participant proposes a specific work up and management plan to correct (or requests this be done by the primary team).
26. If the pH is <7.3 or >7.5, the participant proposes a specific work up and management plan to correct (or requests this be done by the primary team).
27. If the total T4 level is <3 or >30 mg/dL, the participant proposes a specific work up and management plan to correct (or requests this be done by the primary team).
28. If the free T4 level is <0.4 or >5 ng/dL, the participant proposes a specific work up and management plan to correct (or requests this be done by the primary team).
29. If all prerequisites are met, the participant moves on to the clinical exam to determine BD/DNC.
30. If unable to adequately correct metabolic derangements but all other prerequisites are met, the participant moves on to the clinical exam to determine BD/DNC.
31. If the extremities are covered to start the case, the participant uncovers the extremities.
32. Participant tests response to visual stimulation ("blink to threat").
33. Participant keeps hand flat while assessing response to visual threat.
34. Participant tests response to loud auditory stimulation.

© 2025 Daniel S. Harrison, Neil Dhruva, Jenna L. Ford, David M. Greer, Sarah Wahlster, Nikita Chhabra, Nicholas A. Morris

35. Participant tests response to central noxious stimulus.
36. Participant tests response to sternal rub.
37. Participant tests response to nasal tickle.
38. Participant tests response to supraorbital notch pressure.
39. Participant tests response to TMJ pressure.
40. Participant tests response to trapezius squeeze.
41. Participant tests response to noxious stimulus on the cranium.
42. Participant tests response to noxious stimulus below the cranium.
43. Participant tests response to peripheral noxious stimulus in all extremities.
44. Participant tests response to a proximal peripheral stimulus (e.g. arm or thigh pinch) in all extremities.
45. Participant tests response to a distal peripheral noxious stimulus (e.g. forearm or leg pinch or nailbed pressure) in all extremities.
46. Participant tests pupillary responses to bright light bilaterally.
47. Participant tests horizontal oculocephalic reflex.
48. Participant tests vertical oculocephalic reflex
49. Participant stabilizes endotracheal tube while checking OCR.
50. Participant tests oculovestibular reflexes on both ears separately.
51. If there is rupture of the tympanic membrane, participant does NOT perform the OVR test.
52. If cerumen impaction is present, the participant either clears the impaction or asks for assistance in doing so.
53. If the head of bed is NOT at 30 degrees to start the OVR test, the participant positions the head of bed at 30 degrees.
54. Participant instills 50cc of ice cold water against the tympanic membrane.

© 2025 Daniel S. Harrison, Neil Dhruva, Jenna L. Ford, David M. Greer, Sarah Wahlster, Nikita Chhabra, Nicholas A. Morris

55. Participant instills ice water against the tympanic membrane over 60 seconds.
56. Participant holds the eyelids open and observes for eye movement for at least 60 seconds while instilling water against the tympanic membrane.
57. Participant waits at least 5 minutes before testing the contralateral ear.
58. Participant tests the corneal reflex bilaterally.
59. Participant touches the cornea at the border of the iris.
60. Participant applies light pressure to the cornea with a cotton swab on a stick.
61. Participant assesses for a gag reflex.
62. Participant uses a tongue depressor, cotton-tipped applicator, or rigid suction device to assess the gag reflex.
63. Participant stimulates the posterior pharyngeal wall bilaterally to assess the gag reflex.
64. Participant assesses for a cough reflex.
65. Participant checks for a jaw jerk reflex.
66. The participant proceeds to the apnea test if the clinical exam is consistent with BD/DNC.
67. If there is concern for a high cervical spinal cord injury, the participant does NOT perform an apnea test.
68. Participant asks the primary team about or personally assesses the patient’s volume status.
69. If the patient is found to be hypovolemic, the participant administers fluids or asks the primary team to administer fluids.
70. If an arterial line is NOT already in place, participant requests placement of arterial line or places it themself.
71. Participant asks for telemetry to be displayed on monitor.
72. Participant asks for blood pressure to be displayed on monitor.
73. If unable to obtain arterial access, participant asks for frequent cycling of BP cuff.

© 2025 Daniel S. Harrison, Neil Dhruva, Jenna L. Ford, David M. Greer, Sarah Wahlster, Nikita Chhabra, Nicholas A. Morris

74. Participant asks for oxygen saturation to be displayed on monitor.
75. If performing apnea test on a ventilator, participant asks for end tidal CO2 to be displayed on monitor.
76. Participant requests presence of respiratory therapist.
77. If the nurse asks to leave the room, the participant requests that the nurse remain available for the apnea test.
78. If participant does NOT have appropriate expertise in managing cardiopulmonary complications, they request presence of staff with appropriate expertise in managing cardiopulmonary complications.
79. If necessary supplies for apnea testing are NOT present, the participant requests these.
80. If blood gas syringes are NOT present, the participant requests these.
81. If vasopressors are NOT present, the participant requests these.
82. If inotropes are NOT present, the participant requests these.
83. If IV fluids are NOT present, the participant requests these.
84. Participant requests pre-oxygenation for at least 10 minutes with 100% FiO2.
85. If PEEP >5, participant requests decreasing PEEP to 5, if the patient can tolerate.
86. Participant requests baseline ABG.
87. If the patient is on VA ECMO, participant instructs the embedded simulation participant (ESP) to draw ABGs peripherally and from ECMO post-circuit oxygenator.
88. If PaO2 <200, then participant requests pre-oxygenation at a higher PEEP and repeat ABG.
89. If unable to achieve adequate pre-oxygenation despite PEEP titration, the participant does NOT perform an apnea test.
90. If pH <7.35 or >7.45, then participant makes changes or requests that the primary team make changes in ventilator settings to achieve this.

© 2025 Daniel S. Harrison, Neil Dhruva, Jenna L. Ford, David M. Greer, Sarah Wahlster, Nikita Chhabra, Nicholas A. Morris

91. If unable to achieve pH 7.35-7.45, the participant does NOT perform an apnea test.
92. If PaCO2 <35 or >45 and the patient does NOT have known chronic hypercarbia, then participant makes changes or requests that the primary team make changes in ventilator settings and rechecks an ABG prior to disconnecting the patient from the ventilator.
93. If the patient has known chronic hypercarbia and is NOT at their known or estimated baseline PaCO2, the participant makes changes or requests that the primary team make changes in ventilator settings and rechecks an ABG prior to disconnecting the patient from the ventilator.
94. If patient requires pre-oxygenation at PEEP >5, then participant performs a modified rather than conventional apnea test.
95. (Only allowed when criteria for conventional apnea testing are met) Participant places or instructs RT to place a catheter with diameter <70% of the tracheal tube into the tracheal tube.
96. (Only allowed when criteria for conventional apnea testing are met) Participant instructs Embedded Simulation Participant (ESP) to deliver 100% FiO2.
97. (Only allowed when criteria for conventional apnea testing are met) Participant instructs ESP to deliver oxygen at a flow rate of 4-6L/min
98. (Allowed regardless of whether criteria for conventional apnea testing are met) Participant instructs RT to use flow inflating resuscitation bag or t-piece with functioning PEEP valve; OR Participant instructs RT to switch ventilator to CPAP mode and disables apnea ventilation, silences alarm, removes condensation from circuit, positions circuit away from the patient's body, and asks that the trigger sensitivity be set to avoid auto-triggering and is sensitive enough to detect a breath.
99. Participant asks for the chest and abdomen to be uncovered.
100. Participant palpates the chest to assess for movement.
101. If using a flow-inflating bag, participant watches and feels the bag.

© 2025 Daniel S. Harrison, Neil Dhruva, Jenna L. Ford, David M. Greer, Sarah Wahlster, Nikita Chhabra, Nicholas A. Morris

102. If using the ventilator, participant watches the ventilator monitor to assess for spontaneous breathing.
103. Participant requests transcutaneous $CO_2$ monitoring.
104. Participant checks an ABG after 8-10 min of apnea.
105. If the patient develops hypotension or oxygen desaturation and the need to abort the apnea test seems imminent, the participant obtains an ABG as they are placing the patient back on the ventilator.
106. If the $PaCO_2$ and pH level criteria are NOT reached, and the patient did NOT experience hemodynamic instability or hypoxemia during apnea testing, the participant either continues the apnea test beyond 10 minutes with ABG measurements checked at least every 2 minutes; OR repeats apnea testing for a longer period after again preoxygenating and reestablishing baseline $PaCO_2$ and pH levels.
107. If there is spontaneous respiratory effort, the participant reconnects the ventilator and does NOT continue with further BD/DNC evaluation.
108. If there is hemodynamic instability (MAP <75 or SBP <100 despite attempted correction with vasopressors, inotropes, or fluids), the participant reconnects the ventilator.
109. If there is hypoxia ($O_2$ sat <85%), the participant reconnects the ventilator.
110. If there is arrhythmia with hemodynamic instability, the participant reconnects the ventilator.
111. If the pH is <7.3 and $pCO_2$ is >60 and >20 above the baseline, the participant reconnects the ventilator.
112. The participant adjusts (or requests primary team to adjust) ventilator settings to achieve normoxia, normocapnea, and normal acid-base status, after the apnea test.

© 2025 Daniel S. Harrison, Neil Dhruva, Jenna L. Ford, David M. Greer, Sarah Wahlster, Nikita Chhabra, Nicholas A. Morris

113. If conventional apnea testing is performed, participant performs or requests a recruitment maneuver (positive end-expiratory pressure of 15 cm H20 for 15 seconds, then 30cm H20 for 30 seconds) after reconnecting the patient to the ventilator.
114. If evaluation is consistent with BD/DNC, participant declares BD/DNC.
115. If ancillary testing is performed, the participant correctly determines the time of death after ancillary testing as the time attending clinician documents in the medical record that the ancillary test results are consistent with BD/DNC.
116. If unable to correct metabolic derangements, the participant requests an ancillary test.
117. If patient's baseline CO2 is unknown, the participant requests an ancillary test.
118. If unable to perform apnea test (e.g. due to concern for high cervical spinal injury or hypoxemia), the participant requests an ancillary test.
119. If required by hospital/state guidelines, the participant requests an ancillary test.
120. If an ancillary test is required, participant requests diagnostic cerebral angiogram, transcranial Doppler ultrasound (adult patients only), or radionuclide perfusion scintigraphy.
121. If it was unclear whether an observed limb movement was spinally- (vs. cerebrally-) mediated, the participant asks for an ancillary test.
122. If a portion of the clinical exam could NOT be completed (aside from the oculocephalic reflex) or is confounded, the participant asks for an ancillary test.
123. If anophthalmia is present, the participant asks for an ancillary test.
124. If corneal trauma or transplantation is present, the participant asks for an ancillary test.
125. If fracture of the base of the skull or petrous temporal bone is present, the participant asks for an ancillary test.
126. If there is concern for a high cervical spinal cord injury, the participant asks for an ancillary test.

127. If there is a history of ophthalmic surgery that influences pupillary reactivity, the participant asks for an ancillary test.
128. If there is severe facial trauma, the participant asks for an ancillary test.
129. If there is history of a severe pre-existing neuromuscular disorder, the participant asks for an ancillary test.
130. If there is severe orbital or scleral edema or chemosis, the participant asks for an ancillary test.

*Panel proposed*

1. If prior neuroimaging is NOT consistent with mechanism and severity of brain injury or does NOT demonstrate catastrophic, permanent supratentorial injury, then participant requests repeat neuroimaging before performing a clinical exam to determine BD/DNC.
2. If insufficient time has passed since medical or surgical interventions to treat elevated intracranial pressure to ensure there is no potential for recovery of brain function as determined by the evaluator based on the expected timeline of the intervention, participant does NOT perform a clinical exam to determine BD/DNC.
3. For children only, if BP is NOT at age-appropriate goal, participant asks for BP augmentation before performing (or continuing with) a clinical exam to determine BD/DNC.
4. If apnea is suspected but patient-triggered breaths are noted on the ventilator, then participant proposes a specific work up for auto-triggering/cycling (or requests this be done by the primary team).
5. If there is concern for a cervical spinal injury or skull base injury, the participant does NOT test oculocephalic reflexes.
6. (Allowed regardless of whether criteria for conventional apnea testing are met) Participant instructs RT to switch ventilator to CPAP mode and disables apnea ventilation, silences alarm, removes condensation from circuit, positions circuit away from the patient's body, and asks that

© 2025 Daniel S. Harrison, Neil Dhruva, Jenna L. Ford, David M. Greer, Sarah Wahlster, Nikita Chhabra, Nicholas A. Morris

the trigger sensitivity be set to avoid auto-triggering and is sensitive enough to detect a breath. *Please note that this is only one of several possibly appropriate techniques for apnea testing. Please vote on the importance of this item if the participant has chosen to perform the apnea test using CPAP.*

7. If using a flow-inflating bag, participant feels the bag
8. Participant checks an ABG after 5 min of apnea.
