## Supplementary material for "A Modified Delphi Consensus-based Comprehensive Checklist and Angoff Standard for Assessment of Competency in Brain Death/Death by Neurologic Criteria Determination": eAppendix 2

**eAppendix 2- Summary of Steering Committee Modifications to Final Checklist**

**Combined items**
*Panelist retained items*
(For adult patients only, if the participant has chosen to perform apnea testing with tracheal insufflation) Participant instructs Embedded Simulation Participant (ESP) to deliver 100% FiO2.
(For adult patients only, if the participant has chosen to perform apnea testing with tracheal insufflation) Participant instructs the Embedded Simulation Participant to deliver oxygen at a flow rate of 4-6L/min.
*Steering committee combined item*
(For adult patients only, if the participant has chosen to perform apnea testing with tracheal insufflation) Participant instructs Embedded Simulation Participant (ESP) to deliver 100% FiO2 at a flow rate of 4-6 L/min.
*Rationale*
Supplemental O2 provided through a catheter is delivered at 100% FiO2 by convention.

**Modified items**
*Panelist retained item*
If clinical exam and apnea test are consistent with BD/DNC and there is no indication for ancillary testing, then participant declares BD/DNC.
*Steering committee modified item*
If clinical exam, apnea test, and ancillary testing (if indicated) are consistent with BD/DNC, then participant declares DB/DNC.
*Rationale*
Accurate determination of brain death is important regardless of whether ancillary testing is indicated.

*Panelist retained item*
If ancillary testing is performed, the participant correctly determines the time of death after ancillary testing as the time attending clinician documents in the medical record that the ancillary test results are consistent with BD/DNC.
*Steering committee modified item*
The participant correctly determines the time of death.
*Rationale*
Correctly determining the time of death is important regardless of whether ancillary testing is indicated.

**Removed items**
Participant tests response to peripheral noxious stimulus in all extremities.
*Rationale*
The following more specific items were retained:
Participant tests response to a proximal peripheral stimulus (e.g. arm or thigh pinch) in all extremities.
Participant tests response to a distal peripheral noxious stimulus (e.g. forearm or leg pinch or nailbed pressure) in all extremities.

Participant tests response to central noxious stimulus.
*Rationale*

The following more specific items were retained:
Participant tests response to noxious stimulus on the cranium.
Participant tests response to noxious stimulus below the cranium.

Participant instructs RT to use flow inflating resuscitation bag or t-piece with functioning PEEP valve.
*Rationale*
This item was included in the modified Delphi process to assess whether panelists felt use of flow inflating resuscitation bag or t-piece with functioning PEEP valve was an appropriate technique for apnea testing but was not intended to be a final checklist item.

**Added items**
"The Delphi panel voted to retain 3 techniques for maintaining adequate oxygenation during apnea testing:
1. Use of tracheal insufflation (for adult patients only)
2. Use of flow inflating resuscitation bag or t-piece with functioning PEEP valve
3. Use of CPAP on the ventilator
Please indicate the percentage of minimally competent participants you would expect to perform apnea testing using any of the above techniques."
*Rationale*
Delphi panelists voted to retain items related to each of the above techniques.

Participant performs otoscopy bilaterally prior to checking the OVR
*This item only applies when a task trainer for otoscopy is available*
*Rationale*
The final checklist included an item for clearing cerumen impaction but not for identifying it.
