## Supplementary material for "A Modified Delphi Consensus-based Comprehensive Checklist and Angoff Standard for Assessment of Competency in Brain Death/Death by Neurologic Criteria Determination": eTable 2

eTable 2- Modifiable checklist for facilitator or rater

| Checklist Item | Complete? |
| --- | --- |
| Participant does NOT perform a clinical exam to determine BD/DNC if unable to confirm a catastrophic, permanent brain injury caused by an identified mechanism that is known to lead to BD/DNC. |  |
| Comments: |  |
| Participant reviews neuroimaging studies. |  |
| Comments: |  |
| If prior neuroimaging is NOT consistent with mechanism and severity of brain injury or does NOT demonstrate catastrophic, permanent supratentorial injury, then participant requests repeat neuroimaging before performing a clinical exam to determine BD/DNC. |  |
| Comments: |  |
| If insufficient time has passed since medical or surgical interventions to treat elevated intracranial pressure to ensure there is no potential for recovery of brain function as determined by the evaluator based on the expected timeline of the intervention, participant does NOT perform a clinical exam to determine BD/DNC. |  |
| Comments: |  |
| If patient core body temperature is or has been < or = 35.5, participant requests rewarming to > 36C for >24 hours before performing a clinical exam to determine BD/DNC. |  |
| Comments: |  |
| For adult patients only, if SBP < 100 (in a patient NOT on veno-arterial ECMO) or MAP < 75, participant asks for BP augmentation before performing (or continuing with) a clinical exam to determine BD/DNC. |  |
| Comments: |  |
| For children only, if BP is NOT at age-appropriate goal, participant asks for BP augmentation before performing (or continuing with) a clinical exam to determine BD/DNC. |  |
| Comments: |  |
| Participant asks about or reviews recently administered medications. |  |
| Comments: |  |
| If a paralytic has been given, participant uses train-of-four stimulator or demonstrates deep tendon reflexes to exclude ongoing paralysis. |  |
| Comments: |  |
| If a paralytic has been given and DTRs are absent or there are no twitches on TOF, then participant asks to wait and reassess until one or both of these return before performing a clinical exam to determine BD/DNC. |  |
| Comments: |  |
| Participant asks about or reviews drug levels, as available, for medications that may suppress central nervous system function. |  |
| Comments: |  |
| If drug levels for medications that may suppress central nervous system function are supratherapeutic, pentobarbital level is >5mcg/mL, blood or urine toxicology screen is positive, or blood alcohol level is >80m/dL, then participant asks to recheck the level or screen after a delay before performing a clinical exam to determine BD/DNC. |  |
| Comments: |  |
| If the patient received CNS-suppressing medications for which levels are NOT available, the participant asks to wait at least five half-lives for these drugs to have passed before performing a clinical exam to determine BD/DNC. |  |
| Comments: |  |
| If the patient received CNS-suppressing medications for which levels are NOT available and there is renal/hepatic dysfunction or if the patient is obese or was hypothermic, the participant asks to wait longer than five half-lives before performing a clinical exam to determine BD/DNC. |  |
| Comments: |  |
| If apnea is suspected but assisted breaths are noted on the ventilator, then participant assesses for auto-triggering/cycling by the ventilator (or requests this be done by the primary team). |  |
| Comments: |  |
| The participant reviews or requests results of recent laboratory values. |  |
| Comments: |  |
| If a severe metabolic, acid-base, or endocrine derangement is present and correctable, the participant proposes a specific work up and management plan to correct (or requests this be done by the primary team), if correction with reasonable interventions is feasible. |  |
| Comments: |  |
| If the glucose level is < 70 or > 300 mg/dL, the participant proposes a specific work up and management plan to correct (or requests this be done by the primary team), if correction with reasonable interventions is feasible. |  |
| Comments: |  |
| If the magnesium level is < 1.5 or > 4 mg/dL, the participant proposes a specific work up and management plan to correct (or requests this be done by the primary team), if correction with reasonable interventions is feasible. |  |
| Comments: |  |
| If the potassium level is < 3 or > 6 mmol/L, the participant proposes a specific work up and management plan to correct (or requests this be done by the primary team), if correction with reasonable interventions is feasible. |  |
| Comments: |  |
| If the pH is < 7.3 or > 7.5, the participant proposes a specific work up and management plan to correct (or requests this be done by the primary team), if correction with reasonable interventions is feasible. |  |
| Comments: |  |
| If all prerequisites are met, the participant moves on to the clinical exam to determine BD/DNC. |  |
| Comments: |  |
| If unable to adequately correct metabolic derangements but all other prerequisites are met, the participant moves on to the clinical exam to determine BD/DNC. |  |
| Comments: |  |
| If the extremities are covered to start the case, the participant uncovers the extremities. |  |
| Comments: |  |
| Participant tests response to visual stimulation ("blink to threat"). |  |
| Comments: |  |
| Participant tests response to loud auditory stimulation. |  |
| Comments: |  |
| Participant tests response to noxious stimulus on the cranium. |  |
| Comments: |  |
| Participant tests response to supraorbital notch pressure. |  |
| Comments: |  |
| Participant tests response to noxious stimulus below the cranium. |  |
| Comments: |  |
| Participant tests response to a proximal peripheral stimulus (e.g. arm or thigh pinch) in all extremities. |  |
| Comments: |  |
| Participant tests response to a distal peripheral noxious stimulus (e.g. forearm or leg pinch or nailbed pressure) in all extremities. |  |
| Comments: |  |
| Participant tests pupillary responses to bright light bilaterally. |  |
| Comments: |  |
| Participant tests horizontal oculocephalic reflex. |  |
| Comments: |  |
| Participant stabilizes endotracheal tube while checking OCR. |  |
| Comments: |  |
| If there is concern for a cervical spinal injury or skull base injury, the participant does NOT test oculocephalic reflexes. |  |
| Comments: |  |
| Participant tests oculovestibular reflexes on both ears separately. *This item only applies when a task trainer for assessment of the OVR is available. When a task trainer is unavailable, the participant must verbally describe this item.* |  |
| Comments: |  |
| Participant performs otoscopy bilaterally prior to checking the OVRs *This item only applies when a task trainer for assessment of the OVR is available. When a task trainer is unavailable, the participant must verbally describe this item.* |  |
| Comments: |  |
| If cerumen impaction is present, the participant either clears the impaction or asks for assistance in doing so. *This item only applies when a task trainer for assessment of the OVR is available. When a task trainer is unavailable, the participant must verbally describe this item.* |  |
| Comments: |  |
| If the head of bed is NOT at 30 degrees to start the OVR test, the participant positions the head of bed at 30 degrees. *This item only applies when a task trainer for assessment of the OVR is available. When a task trainer is unavailable, the participant must verbally describe this item.* |  |
| Comments: |  |
| Participant instills 50cc of ice cold water against the tympanic membrane. *This item only applies when a task trainer for assessment of the OVR is available. When a task trainer is unavailable, the participant must verbally describe this item.* |  |
| Comments: |  |
| Participant instills ice water against the tympanic membrane over 60 seconds. *This item only applies when a task trainer for assessment of the OVR is available. When a task trainer is unavailable, the participant must verbally describe this item.* |  |
| Comments: |  |
| Participant holds the eyelids open and observes for eye movement for at least 60 seconds while instilling water against the tympanic membrane. *This item only applies when a task trainer for assessment of the OVR is available. When a task trainer is unavailable, the participant must verbally describe this item.* |  |
| Comments: |  |
| Participant waits at least 5 minutes before testing the contralateral ear. *This item only applies when a task trainer for assessment of the OVR is available. When a task trainer is unavailable, the participant must verbally describe this item.* |  |
| Comments: |  |
| Participant tests the corneal reflex bilaterally. |  |
| Comments: |  |
| Participant touches the cornea at the border of the iris. |  |
| Comments: |  |
| Participant applies light pressure to the cornea with a cotton swab on a stick. |  |
| Comments: |  |
| Participant assesses for a gag reflex. |  |
| Comments: |  |
| Participant uses a tongue depressor, cotton-tipped applicator, or rigid suction device to assess the gag reflex. |  |
| Comments: |  |
| Participant stimulates the posterior pharyngeal wall bilaterally to assess the gag reflex. |  |
| Comments: |  |
| Participant assesses for a cough reflex. |  |
| Comments: |  |
| The participant proceeds to the evaluation of safety and appropriateness of the apnea test if the clinical exam is consistent with BD/DNC. |  |
| Comments: |  |
| Participant asks the primary team about or personally assesses the patient’s volume status. |  |
| Comments: |  |
| If the patient is found to be hypovolemic, the participant administers fluids or asks the primary team to administer fluids. |  |
| Comments: |  |
| Participant asks for 3 lead ECG to be displayed on monitor. |  |
| Comments: |  |
| Participant asks for blood pressure to be displayed on monitor. |  |
| Comments: |  |
| If unable to obtain arterial access, participant asks for frequent cycling of BP cuff. |  |
| Comments: |  |
| Participant asks for oxygen saturation to be displayed on monitor. |  |
| Comments: |  |
| Participant requests presence of respiratory therapist (if required by local protocol). |  |
| Comments: |  |
| If participant does NOT have appropriate expertise in managing cardiopulmonary complications, they request presence of staff with appropriate expertise in managing cardiopulmonary complications. |  |
| Comments: |  |
| If necessary supplies for apnea testing are NOT present, the participant requests these. |  |
| Comments: |  |
| If blood gas syringes are NOT present, the participant requests these. |  |
| Comments: |  |
| If vasopressors are NOT present, the participant requests these. |  |
| Comments: |  |
| If IV fluids are NOT present, the participant requests these. |  |
| Comments: |  |
| Participant requests pre-oxygenation for at least 10 minutes with 100% FiO2. |  |
| Comments: |  |
| Participant requests baseline ABG. |  |
| Comments: |  |
| If the patient is on VA ECMO, participant instructs the embedded simulation participant (ESP) to draw ABGs peripherally and from ECMO post-circuit oxygenator. |  |
| Comments: |  |
| If pH < 7.35 or > 7.45, then participant makes changes or requests that the primary team make changes in ventilator settings to achieve this. |  |
| Comments: |  |
| If PaCO2 < 35 or > 45 and the patient does NOT have known chronic hypercarbia, then participant makes changes or requests that the primary team make changes in ventilator settings and rechecks an ABG prior to disconnecting the patient from the ventilator. |  |
| Comments: |  |
| If the patient has known chronic hypercarbia and is NOT at their known or estimated baseline PaCO2, the participant makes changes or requests that the primary team make changes in ventilator settings and rechecks an ABG prior to disconnecting the patient from the ventilator. |  |
| Comments: |  |
| The participant performs apnea testing using any of the below techniques for maintaining adequate oxygenation: 1. Use of tracheal insufflation (for adult patients only) 2. Use of flow inflating resuscitation bag or t-piece with functioning PEEP valve 3. Use of CPAP on the ventilator |  |
| Comments: |  |
| (For adult patients only, if the participant has chosen to perform apnea testing with tracheal insufflation) Participant places or instructs RT to place a catheter with diameter < 70% of the tracheal tube into the tracheal tube. |  |
| Comments: |  |
| (For adult patients only, if the participant has chosen to perform apnea testing with tracheal insufflation) Participant instructs Embedded Simulation Participant (ESP) to deliver 100% FiO2 at a flow rate of 4-6 L/min. |  |
| Comments: |  |
| Participant asks for the chest and abdomen to be uncovered. |  |
| Comments: |  |
| If using a flow-inflating bag, participant watches the bag. |  |
| Comments: |  |
| If the participant has chosen to perform the apnea test on the ventilator using CPAP, the participant instructs RT to switch ventilator to CPAP mode and disables default back up apnea ventilation, silences alarm, removes condensation from circuit, positions circuit away from the patient's body, and asks that the trigger sensitivity be set to avoid auto-triggering and is sensitive enough to detect a true spontaneous respiratory effort. |  |
| Comments: |  |
| If using the ventilator, participant watches the ventilator monitor to assess for spontaneous breathing. |  |
| Comments: |  |
| Participant checks an ABG after 8-10 min of apnea. |  |
| Comments: |  |
| If the patient develops hypotension or oxygen desaturation and the need to abort the apnea test seems imminent, the participant obtains an ABG as they are placing the patient back on the ventilator. |  |
| Comments: |  |
| If the PaCO2 and pH level criteria are NOT reached and the patient did NOT experience hemodynamic instability or hypoxemia during apnea testing, the participant either continues the apnea test beyond 10 minutes with repeat ABG measurements; OR repeats apnea testing for a longer period after again preoxygenating and reestablishing baseline PaCO2 and pH levels. |  |
| Comments: |  |
| If there is spontaneous respiratory effort, the participant reconnects the ventilator and does NOT continue with further BD/DNC evaluation. |  |
| Comments: |  |
| If there is hemodynamic instability (MAP < 75 or SBP < 100 despite attempted correction with vasopressors, inotropes, or fluids), the participant reconnects the ventilator. |  |
| Comments: |  |
| If there is arrhythmia with hemodynamic instability, the participant reconnects the ventilator. |  |
| Comments: |  |
| If there is hypoxia (O2 sat < 85%), the participant reconnects the ventilator. |  |
| Comments: |  |
| If the PCO2 is > 60 and > 20 above the baseline, the participant reconnects the ventilator. |  |
| Comments: |  |
| The participant adjusts (or requests primary team to adjust) ventilator settings to achieve normoxia, normocapnea, and normal acid-base status after the apnea test. |  |
| Comments: |  |
| If clinical exam, apnea test, and ancillary testing (if indicated) are consistent with BD/DNC, then participant declares DB/DNC. |  |
| Comments: |  |
| The participant correctly determines the time of death. |  |
| Comments: |  |
| If unable to correct metabolic derangements, the participant requests an ancillary test. |  |
| Comments: |  |
| If unable to perform apnea test (e.g. due to concern for high cervical spinal injury or hypoxemia), the participant requests an ancillary test. |  |
| Comments: |  |
| If required by hospital/state guidelines, the participant requests an ancillary test. |  |
| Comments: |  |
| If an ancillary test is required, participant requests diagnostic cerebral angiogram, transcranial Doppler ultrasound (adult patients only), or radionuclide perfusion scintigraphy. |  |
| Comments: |  |
| If it was unclear whether an observed limb movement was spinally- (vs. cerebrally-) mediated, the participant asks for an ancillary test. |  |
| Comments: |  |
| If a portion of the clinical exam could NOT be completed (aside from the oculocephalic reflex) or is confounded, the participant asks for an ancillary test. |  |
| Comments: |  |
| If anophthalmia is present, the participant asks for an ancillary test. |  |
| Comments: |  |
| If corneal trauma or transplantation is present, the participant asks for an ancillary test. |  |
| Comments: |  |
| If there is concern for a high cervical spinal cord injury, the participant asks for an ancillary test. |  |
| Comments: |  |
| If there is a history of ophthalmic surgery that influences pupillary reactivity, the participant asks for an ancillary test. |  |
| Comments: |  |
| If there is history of a severe pre-existing neuromuscular disorder, the participant asks for an ancillary test. |  |
| Comments: |  |
